# In Utero Exposure to the Great Depression is Reflected in Late-Life Epigenetic Aging Signatures

**DOI:** 10.1101/2022.05.18.22275258

**Authors:** Lauren L. Schmitz, Valentina Duque

## Abstract

Research on maternal-fetal epigenetic programming argues that adverse exposures to the intrauterine environment can have long-term effects on adult morbidity and mortality. However, causal research on epigenetic programming in humans at a population level is rare and is often unable to separate intrauterine effects from conditions in the postnatal period that may continue to impact child development. In this study, we used a quasi-natural experiment that leverages state-year variation in economic shocks during the Great Depression to examine the causal effect of environmental exposures in early life on late-life accelerated epigenetic aging for 832 participants in the U.S. Health and Retirement Study (HRS). HRS is the first population-representative study to collect epigenome-wide DNA methylation data that has the sample size and geographic variation necessary to exploit quasi-random variation in environments, which expands possibilities for causal research in epigenetics. Our findings suggest that exposure to changing economic conditions in the 1930s had lasting impacts on next-generation epigenetic aging signatures that were developed to predict mortality risk (GrimAge) and physiological decline (DunedinPoAm). We show that these effects are localized to the in utero period specifically as opposed to the pre-conception, postnatal, childhood, or early adolescent periods. After evaluating changes in mortality rates for Depression-era birth cohorts, we conclude that these effects likely represent lower bound estimates of the true impacts of the economic shock on long-term epigenetic aging.

## Introduction

Aging is characterized by the gradual accumulation of cellular damage, leading to physiological deterioration, loss of function, and increased vulnerability to death (1). A growing body of research suggests that the beginnings of human aging occur during the initial phases of embryogenesis—a paradigm shift in aging research that anchors the onset of age-related damage accumulation to the prenatal period as opposed to later in the life course after the completion of development and the onset of reproductive age (2, 3). The idea that aging later in life could be linked to in utero programming has important implications for the development of early-life interventions that could postpone age-related morbidity and mortality and significantly extend healthy life span (2).

Early-life programming of aging and longevity has its roots in the developmental origins of health and disease (DOHaD) work of David Barker (4, 5), and in the field of environmental epigenetics, which linked the molecular basis for this mode of inheritance to epigenetic mechanisms in animal models (6). Recent quantitative tests of this hypothesis (7, 8) have been possible due to discoveries in epigenetic profiling and machine learning technologies that led to the development of epigenetic aging measures or “epigenetic clocks” that can more accurately track biological age in utero and across the life course (9–21). While age-related deterioration and damage is reflected in nearly every biological process at the molecular and cellular level, the dysregulation of these processes is a function of upstream epigenomic changes that control transcriptional and chromatin networks. In humans, the most well-researched type of epigenetic modification is DNA methylation (DNAm), which refers to the addition of a methyl group to a cytosine nucleotide at cytosine-phosphate-guanine (CpG) site. Epigenetic clocks are calculated by taking the genome-wide weighted average of DNAm levels at CpG sites that are highly associated with either chronological age (first generation clocks) or phenotypic hallmarks of aging (second generation clocks). Epigenetic clocks accurately predict chronological age (9, 17–19, 22, 23), and numerous studies have linked deviations between DNAm age and chronological age— i.e., epigenetic age acceleration (EAA)—with age-related diseases and mortality (24–35), suggesting EAA measures may serve as molecular biomarkers of aging that reflect both resilience and vulnerability during the aging process. More recently, pace of aging measures have been developed using the change in 18 age-associated biomarkers in the same cohort of individuals over time, as opposed to cross-sectional measurements in different individuals (16, 36). Second generation clocks and pace of aging measures have shown more consistent associations with socioeconomic disparities across the life course and are more predictive of morbidity and mortality than first generation clocks that were developed to predict chronological age (37–39). Overall, epigenetic aging measures tend to outperform other biomarkers of aging in predicting lifespans (14, 15, 40), and their correlations with age-related conditions makes them useful in a variety of contexts, including anti-aging interventions (13).

However, despite these advances in molecular aging research, existing causal evidence on the long-term impacts of early-life epigenetic programming in humans at a population level is rare. Causal research in the past has been limited to smaller natural experiments, including the Dutch Hunger Winter Families Study (n_treated_=348) (41–43), Project Ice Storm (n_treated_=34) (44–47), or research on Holocaust survivors (n_treated_=54) (48), that examined the impact of maternal nutrition or stress on DNAm changes in pre-selected CpG regions that regulate specific genes. These findings suggested that adverse maternal environments early in human gestation could result in persistent changes in epigenetic information in adulthood, particularly with respect to the regulation of metabolic, immune, or neurological pathways. However, although these studies offer several strengths, they were focused on unique events in history that may have limited external validity. In addition, few studies have looked at the impact of environmental exposures at different time points in childhood to identify critical periods in development where environmental insults could have their greatest impact on long-term epigenetic signatures. For example, researchers were not able to separate the impact of maternal-fetal exposures to famine or severe weather from the impact of related conditions after birth that may have continued to affect children postnatally (e.g., maternal stress or nutrition) (49). The identification of such critical periods is crucial to understanding which investments and interventions would be most cost effective in improving healthy life span (50, 51).

In this study, we used a quasi-natural experiment that leverages state-year variation in economic conditions during the Great Depression to examine the causal effect of environmental exposures in early life on late-life epigenetic aging signatures for participants in the Health and Retirement Study (HRS). HRS is the first population-representative study in the U.S. to collect CpG-level data that has the sample size and geographic variation necessary to exploit quasi-random variation in economic conditions across states and over time, which expands possibilities for causal research in epigenetics. By linking state-year macroeconomic data on wages, employment, and consumption to the first sixteen years of HRS participants’ lives, we can also separately examine the impact of economic conditions experienced in utero from those experienced in childhood and adolescence to fully consider evidence for “critical periods”.

We focus on the Great Depression for several reasons. First and foremost is the magnitude of the exposure. The Great Depression was the most devastating macroeconomic recession in U.S. history: from 1929 to 1933, real output contracted by more than 25%, prices fell by 33%, and the unemployment rate increased from 3.2% to 25%, reaching the highest levels ever documented in the U.S. (52). The extreme nature of the economic shock was a unique failure of the industrial economy that had devastating effects on individuals’ financial and overall well-being (52, 53). Second, at the time, there were few social welfare programs to ameliorate the widespread economic devastation families experienced. The science of prenatal care was in its infancy, and women lacked access to prenatal vitamins or other nutritional supplements that are now considered vital for fetal development, further exacerbating nutritional deprivation and stressful living conditions for expectant mothers and their children. Third, with respect to study design, because the HRS initially surveyed over 12,000 individuals that were born between 1931 and 1941 when the study began in 1992, we can examine epigenetic aging patterns in a relatively large, population-representative sample of surviving cohort members who had their blood drawn in 2016 (n=832). Finally, since economic recessions are a relatively common phenomenon, findings from this study may have greater external validity and applications to other contexts compared to prior causal research on maternal-fetal epigenetic programming that leveraged unique events in modern history.

Our findings suggest that exposure to economic conditions during the Great Depression had lasting impacts on epigenetic aging signatures, and that these effects were salient in both magnitude and statistical significance for in utero exposures only, strongly suggesting the existence of critical periods of development. Results are specific to next-generation epigenetic aging measures, including GrimAge EAA and the DunedinPoAm (Dunedin(P)ace(o)f(A)ging(m)ethylation) measure, both of which incorporate more complex phenotypes into DNAm algorithms that are trained on clinical outcomes and mortality risk (GrimAge) or on the rate of change in system-integrity biomarkers (DunedinPoAm). A one standard deviation (SD) decrease in wages during the 1930s, which is equal to roughly half of the overall decline in wages experienced during the Great Depression, resulted in ∼0.4 SD increase in accelerated epigenetic aging when participants were between the ages of 75 and 84. Analysis of mortality selection in the 1930s HRS birth cohorts revealed that declines in wages during the prenatal period were associated with an increased probability of death prior to epigenetic profiling in 2016. However, our results are consistent after adjusting our estimates for mortality selection using an inverse probability weighted (IPW) estimator (54), indicating the effects of mortality bias are minor overall and if anything may be biasing our effects towards zero since individuals that survived to older ages appear to be positively selected. Finally, we show that compared to other aging and health phenotypes, accelerated GrimAge EAA and DunedinPoAm signatures are more sensitive indicators of both early-life exposure to economic conditions *and* subsequent mortality, which suggests epigenetic aging measures may contain additional valuable information that could further our understanding of the causes of social disparities in aging and healthspan.

## Results

To conduct our analyses, we linked individual level epigenetic aging measures from the HRS that were profiled in 2016 with macroeconomic data at the state- and year-of-birth level (see **SI Appendix Section 1** and **Table S1** for more details on the epigenetic aging measures used in this study). Annual state-level data that document the dynamics of the macroeconomy in the 1920s and 1930s are rare. Our preferred exposure measure is a wage index from the Bureau of Economic Analysis (BEA) because it includes farm and non-farm wages, which better approximates the economic conditions of families with young children in urban and rural areas (55). In addition, the data are available from 1929 to 1956, which allows us to test the impact of wage fluctuations prenatally through adolescence for individuals born in the 1930s. **Figure 1** documents the variation in the wage index across states relative to 1929 that we exploit in our analysis. Relevant summary statistics for the sample are reported in **SI Appendix, Table S2**.

**Figure 1.**
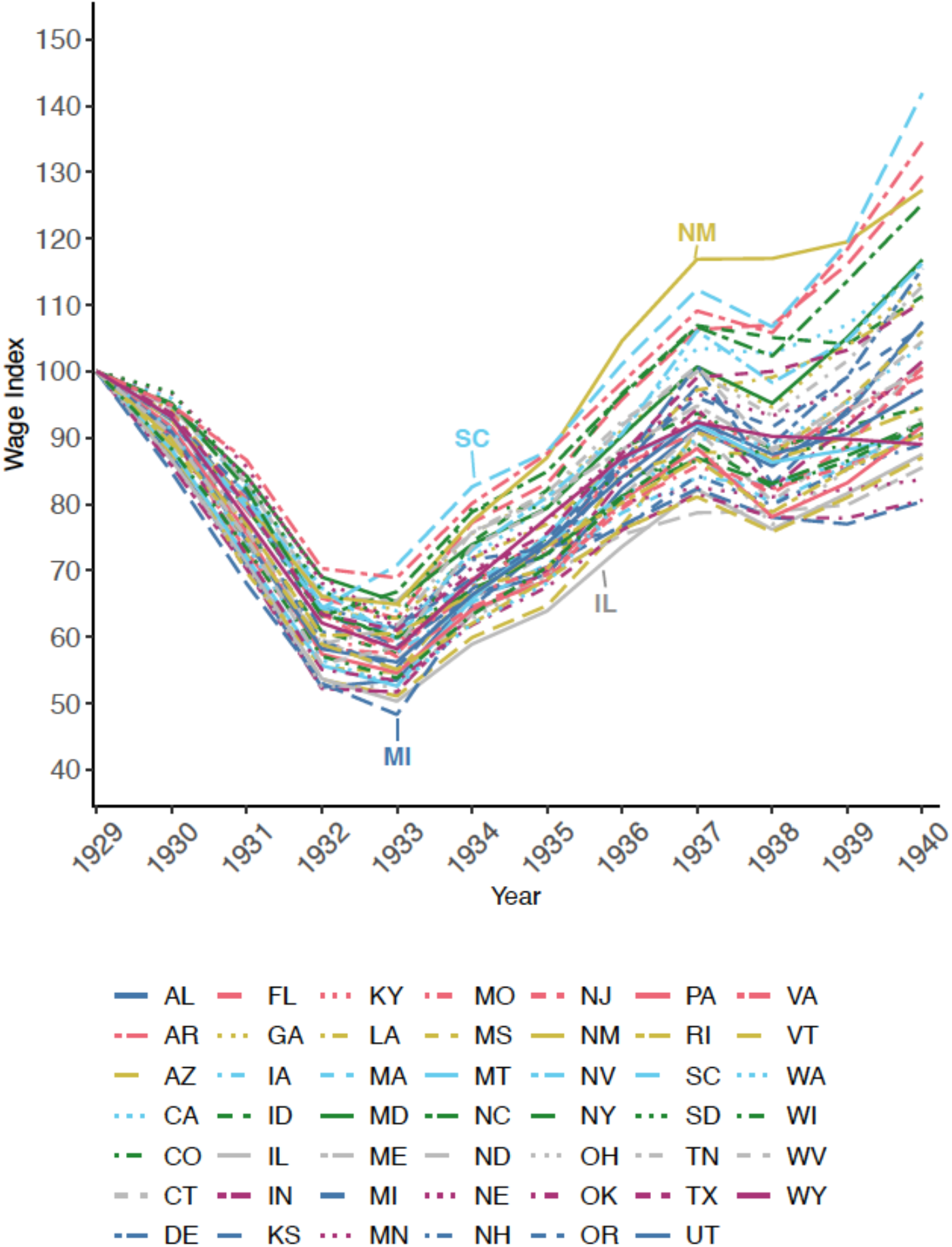
Variation in the wage index across states, 1929-1940 *Notes:* The figure shows fluctuations in unadjusted farm and non-farm wages and salaries relative to 1929. Data were obtained from the Bureau of Economic Analysis (BEA) in index form [SAINC7H Wages and Salaries by Industry (Historical) 1929-1957].

**Table 1** presents results from our baseline specification, which showcases the impact of state-level wages in utero on six epigenetic aging measures constructed from DNAm data profiled in 2016, or when HRS participants were between the ages of 75 and 84. Comparison across multiple epigenetic aging measures is important for the interpretation of our results because each algorithm was developed using different assumptions that capture different aspects of the biological aging process, and corresponding age-adjusted EAA measures are not highly correlated (*r*=0.120-0.605) (**SI Appendix, Figure S1**) (56). Results indicate that variation in wages during the Great Depression had long-term impacts on GrimAge and DunedinPoAm epigenetic aging signatures. A one unit increase in the wage index decreased GrimAge EAA by 0.095 years and decreased the pace or rate of aging as measured by DunedinPoAm by 0.002 years (Bonferroni corrected *p*-value<0.05). To put the magnitude of these effects into perspective, a one standard deviation (SD) decline in wages, or approximately half of the overall decline in wages between 1929 and 1933, resulted in a 0.376 SD increase in GrimAge EAA and a 0.449 SD increase in DunedinPoAm. The magnitude and significance of these results are robust across empirical specifications (**SI Appendix, Table S3**).

**Table 1.** Effect of exposure to the wage index in utero on epigenetic age acceleration (EAA) and pace of aging measures

|  | Horvath<br>EAA | SkinBlood<br>EAA | Hannum<br>EAA | PhenoAge<br>EAA | GrimAge<br>EAA | Dunedin<br>PoAm |
| --- | --- | --- | --- | --- | --- | --- |
| Wage index in utero | 0.0493<br>[0.0647] | -0.0035<br>[0.0579] | -0.0016<br>[0.0700] | 0.0045<br>[0.0670] | -0.0950*<br>[0.0294] | -0.0022*<br>[0.0007] |
| Observations | 832 | 832 | 832 | 832 | 832 | 832 |
| R-squared | 0.15 | 0.142 | 0.189 | 0.17 | 0.273 | 0.134 |
| Effect size per 1 SD increase in<br>the wage index | 0.125 SD | 0.013 SD | 0.005 SD | 0.012 SD | 0.376 SD | 0.449 SD |

*Note:* SD=standard deviation. The table reports the estimated effect of the wage index in utero on EAA and pace of aging measures profiled in 2016. Robust standard errors clustered at the state of birth level in brackets. An asterisk (\*) indicates a significant p-value after Bonferroni correction for 6 independent tests at a family-wise error rate of 0.05 ( $p < 0.0083$ ). All models control for year of birth (YOB) FE, state of birth (SOB) FE, sex, race, and maternal education. State-level controls interacted with YOB linear time trends (LTT) include the 1928 infant mortality rate, the 1929 maternal mortality rate, and whether a state's share of farmland was in the 75th percentile nationally in 1930. The model also includes indicators for whether state employment in manufacturing was in the 75 percentile nationally in 1929 times YOB FE and region of birth-specific LTT. Models were estimated using linear regression with weights provided by the HRS for the Venous Blood Study (VBS) sample.

To determine the extent to which these effects were driven by in utero exposures, we estimated a model that conditioned on exposures to the wage index at different ages, or from the preconception period through age 16. **Figure 2** plots the age-specific exposure coefficients from these specifications for GrimAge EAA and DunedinPoAm. Coefficients are significant for the in utero period only and are similar in magnitude and significance to our baseline results. Importantly, we do not see any evidence of differential trends prior to conception, which is given by the null effect of the wage index two to three years prior to birth, providing support for the identification strategy.

**Figure 2.**
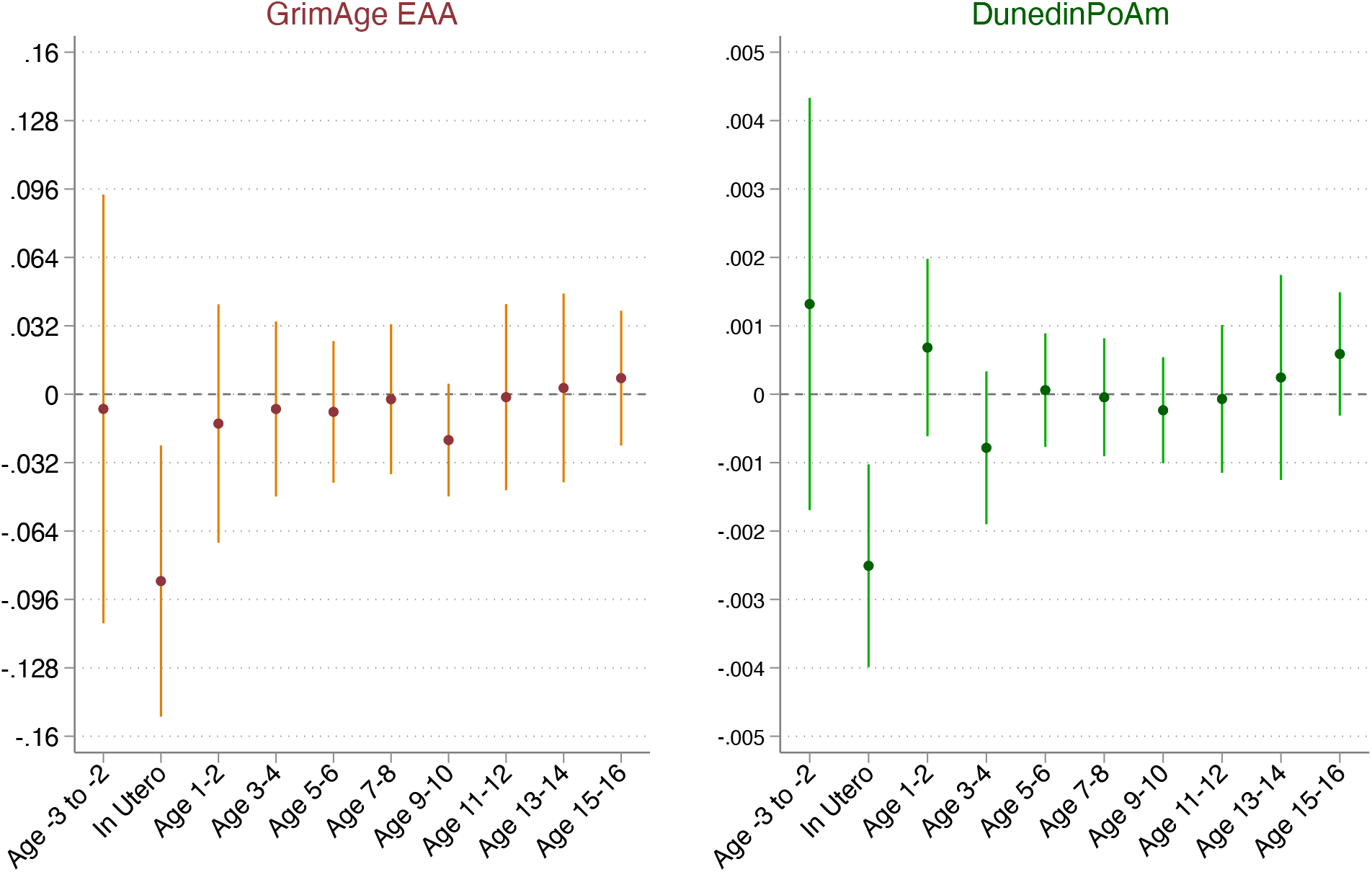
Effect of the wage index during the preconception, in utero, childhood, and adolescent periods on GrimAge EAA and DunedinPoAm *Notes:* EAA=epigenetic age acceleration. The figure reports estimated coefficients from a model that conditions on exposures to the wage index in the preconception, prenatal, childhood, and early adolescent periods for GrimAge EAA and DunedinPoAm as described in Equation (3) (n=832). All models control for year of birth (YOB) FE, state of birth (SOB) FE, sex, race, and maternal education. State-level controls interacted with YOB linear time trends (LTT) include the 1928 infant mortality rate, the 1929 maternal mortality rate, and whether a state’s share of farmland was in the 75th percentile nationally in 1930. The model also includes indicators for whether state employment in manufacturing was in the 75 percentile nationally in 1929 times YOB FE and region of birth-specific LTT. Models were estimated using linear regression with weights provided by the HRS for the Venous Blood Study (VBS) sample. Robust 95% confidence intervals. Standard errors are clustered at the state of birth level.

The salience of exposures during the in utero period is also evident when we use other available state-level data on macroeconomic conditions, including employment and car sales (**SI Appendix, Tables S4-S5**). Employment data reflect labor market fluctuations in manufacturing and non-manufacturing industries (57) and data on car sales proxy household consumption (58). **SI Appendix, Figures S2-S3** depict state-year variation relative to 1929 for the employment and car sales indices. Since these data were not available after 1940, the model is estimated in a subset of individuals with three years of exposure data before birth and two years after birth (n=588). Findings are comparable to results with the wage index. Thus, our findings are not specific to wages but appear to reflect a more consistent pattern between early-life economic conditions and accelerated biological aging.

### Sensitivity analysis

Because DNAm was profiled in whole blood, DNAm measures of aging may reflect differences in the white blood cell (WBC) composition of samples from which the DNA was extracted. Since the relative composition of WBCs changes with age, we tested the sensitivity of our analyses to this variation by adjusting for the percentage of WBC present and their interactions with year of birth fixed effects (**SI Appendix, Table S6**). Adjusting for WBC composition reduced, but did not fully mediate, the magnitude and significance of our results, suggesting our findings may be driven more by extrinsic epigenetic age acceleration (EEAA) as opposed to intrinsic epigenetic age acceleration (IEAA). The EEAA terminology has been used to refer to the observation that clocks trained in whole blood may be more reflective of immune system aging or age-related changes in leukocyte composition that have been more closely linked to metabolic health and environmental stressors, whereas multi-tissue clocks like the Horvath clock that adjust for cell composition are more reflective of cell-intrinsic aging (59, 60). Additionally, results do not appear to be driven by outliers, or individuals in the top and bottom 1% of the GrimAge EAA and DunedinPoAm distributions (**SI Appendix, Table S7**). Finally, because DNAm is in part regulated by genetic polymorphisms, we confirmed that our results are robust to confounding from population stratification in a subsample of European ancestry individuals by adjusting for the first ten principal components of the genetic data and their interaction with the treatment (**SI Appendix, Table S8**).

### Impacts of other co-occurring historical events

The 1930s and ‘40s were a historically rich period characterized by several coinciding events. We analyzed our results in the context of the Dust Bowl, New Deal relief spending, and World War II (WWII) mobilization rates to determine the degree to which they may be influencing our estimates (**SI Appendix, Tables S9-S11**). Overall, our results do not appear to be driven by these co-occurring events, suggesting that economic fluctuations from the Great Depression had an independent effect on biological aging.

### Early-life exposure to economic shocks and mortality

Since DNAm was profiled in our sample at older ages, we conducted additional analyses to understand how mortality selection may be biasing our estimates. While studies have shown that economic conditions in early life can affect mortality (61, 62), evidence on the long-term impacts of the Great Depression on mortality has been mixed (63–66). We re-examined these patterns in the HRS by regressing age-specific survival probabilities on the wage index for respondents born between 1929 and 1940 (n=7,898). Results reveal a positive and significant association between the wage index and the probability of survival from age 75 onwards (**SI Appendix, Table S12**), which overlaps with the age range of our sample in 2016 when epigenetic profiling was conducted (75-84). Additionally, we tested the degree to which survival probabilities vary by mother’s education at baseline (a key proxy of family resources in childhood) (**SI Appendix, Table S12**). We see that survival is positively linked to having a mother with at least a high school degree. Regarding the cause of death, associations between prenatal exposure to the wage index and earlier mortality appear to be driven primarily by deaths from metabolic disorders, which have been linked to intrauterine growth disruptions (**SI Appendix, Table S13**) (5, 41–43). Taken together, these results suggest that our sample is positively selected for survival at older ages.

To investigate the extent that mortality is biasing our estimates, we used fitted values from regression models of survival as inverse probability weights to adjust our estimates so they are more reflective of the HRS sample just prior to the mortality selection (see SI Appendix for details) (54, 67). Survival was modeled using a probit specification under two scenarios: 1) survival until age 75 (the age that we first observe mortality selection in our sample), and 2) survival until 2016 (the year epigenetics were profiled in the HRS). For both scenarios, we present inverse probability weighted (IPW) estimates that use weights constructed with and without adjustments for maternal education in the survival model (**Table 2**). We see some evidence of attenuation bias in our estimates after correcting for mortality selection using the IPW estimator. However, these corrections do not substantially change our estimates, which indicates that any resulting bias from mortality selection may be small and if anything appears to be biasing our estimates downward.

**Table 2.**
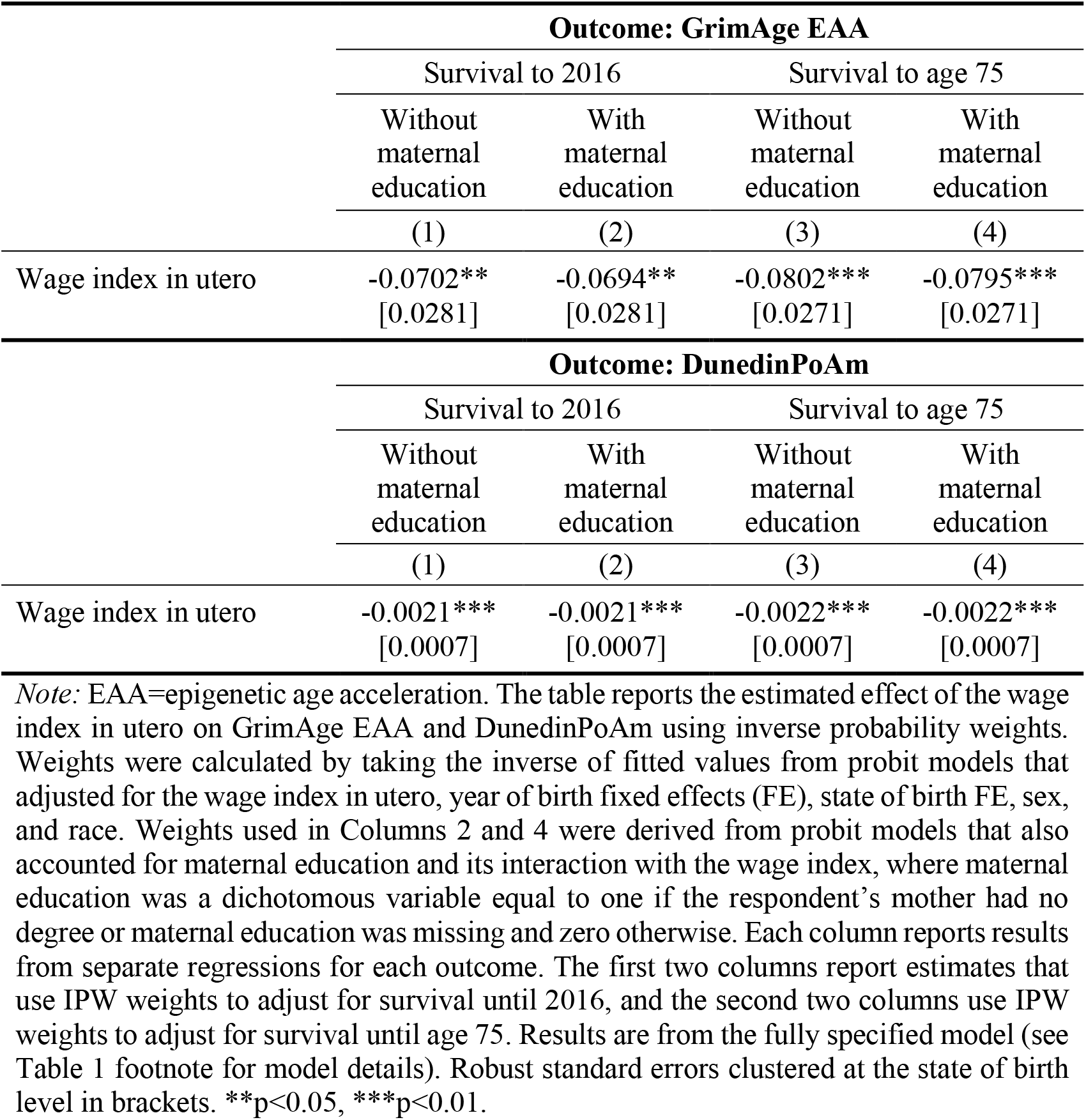
Effect of the wage index in utero on GrimAge EAA and DunedinPoAm, inverse probability weighted (IPW) estimates

### Effects on other aging outcomes and longevity

Finally, we tested whether in utero exposure to wages are predictive of other self-reported or doctor diagnosed measures in our sample, including frailty, metabolic syndrome, self-reported health status (SRHS), and the number of doctor-diagnosed chronic disease conditions (**SI Appendix, Table S14**). These measures are less precise and, apart from the number of chronic disease conditions, the magnitude of their effect sizes (per one SD increase in the wage index) are approximately 40-50% lower relative to GrimAge and DunedinPoAm effect sizes. We then examined the degree to which these measures are associated with the probability of dying in the following HRS wave (approximately 7% of our sample died between 2016 and 2018). GrimAge displays the strongest association in terms of both magnitude (0.173 SD increase in the probability of death per one SD increase in GrimAge) and significance (*p*<0.001), followed by SRHS (0.154 SD; *p*<0.001), DunedinPoAm (0.101 SD; *p*<0.05), and the number of chronic disease conditions (0.077 SD; *p*<0.05) (**SI Appendix, Table S15**). In sum, GrimAge and DunedinPoAm appear to be more sensitive indicators of both in utero exposures to the wage index *and* subsequent longevity than other commonly used measures of aging and health, suggesting they contain additional information on the connection between social disparities in early-life environments and mortality.

## Discussion

In a population-representative sample of over 800 individuals born in the 1930s, we find a significant association between in utero exposure to economic conditions during the Great Depression and late-life epigenetic age acceleration as captured by the GrimAge and DunedinPoAm algorithms. These findings are robust across empirical specifications that account for additional state-level controls and region-specific linear time trends, supporting a causal interpretation. Using the few available sources of state-level variation in macroeconomic conditions from the 1930s, we show that these results are not sensitive to how the economic shock was measured but rather reflect a consistent pattern across changes in wages, consumption, and employment. Although individuals who experienced more favorable economic conditions in early life were more likely to survive to have their epigenetics profiled in 2016, our results were not significantly altered after adjusting for mortality selection in the HRS using an IPW estimator. Thus, any potential bias from the positive selection into our sample that we observe appears to be minimal, and if anything suggests that our estimates represent a lower bound on the effects of economic conditions during the Great Depression on long-term epigenetic aging signatures.

We did not identify any effects for the Horvath, SkinBlood, Hannum, or PhenoAge clocks, which is consistent with prior research that found stronger associations between socioeconomic disadvantage and GrimAge EAA and DunedinPoAm (37–39). Because epigenetic aging measures are composite indicators that are comprised of many different DNAm patterns, a major drawback of their application is a lack of mechanistic understanding of what they are capturing, both in terms of how environmental processes may be initiating these changes as well as their connection to disease etiology (68, 69). Of note, a recent study deconstructed over 5,000 clock CpGs into twelve distinct submodules that display different biological underpinnings and vary considerably in their proportion across clocks (68). GrimAge and DunedinPoAm, which were trained in whole blood to predict mortality or physiological changes with aging, share a very similar composition of submodules that are stronger predictors of mortality and cardiovascular related outcomes. Likewise, while the Horvath, PhenoAge, SkinBlood, and Hannum clocks, which were all trained in some manner on chronological age, are also comprised of mortality-associated modules, they contain additional submodules that have weak or inverse associations with mortality (68). This suggests a connection between our findings and mortality or cardiovascular risk in particular as opposed to tumorigenesis or other age-related cellular processes that appear to be captured more strongly by other clocks.

Overall, it is difficult to disentangle whether the connection between in utero exposures and accelerated biological aging later in life that we observe is operating from 1) epigenetic alterations induced during early development that result in consistently higher incidence of damage throughout life (i.e., fetal programming) (4, 5), or 2) epigenetic signs of aging processes that are accelerated by insults in utero but that continue to develop across the life course (i.e., the idea of high initial damage load or the HIDL hypothesis) (2). In both cases, any downstream consequences of early-life insults will not be readily apparent until we can observe aging at a phenotypic level when progressive accumulation of damage and loss of physiological integrity begin to take hold later in life. Thus, although we cannot use the clocks to disentangle specific mechanistic pathways, we show that they may be particularly sensitive indicators of early-life programming and subsequent mortality risk at later ages. Moreover, this appears to be the case in a subsample of relatively healthier, surviving cohort members that outlived their counterparts. More research is needed, but this indicates that composite measures of epigenetic aging may be especially useful for the detection of disparities in aging prior to the emergence of disease or death.

Along these lines, our findings diverge from prior work on the long-term health effects of the Dust Bowl and the Great Depression that were not able to detect a relationship between in utero exposures and an array of health outcomes in the 1992-2004 HRS waves (66). In part, we hypothesize that differences in the authors’ empirical strategy, which relied on exploiting variation in economic conditions at the region and year of birth level, may have masked substantial heterogeneity in economic activity at the state level that affected the precision of their results. However, effects may also have been biased downward due to the use of self-reported measures that either suffered from measurement error or were not able to detect more subtle differences in aging that were connected to in utero insults.

Some limitations of our study are worth mentioning. First, because monthly macroeconomic data at the state level were not collected in the 1930s, we were not able to identify specific time windows or trimesters during pregnancy that may have been especially vulnerable to the shock, or whether the timing of the exposure mapped onto distinctive epigenetic changes. Second, we cannot identify why and how aggregate economic exposures were affecting the fetal environment, or if our results were driven more by nutritional deprivation, maternal stress, fewer economic resources, or a combination of these factors. Future research will be better poised to address these limitations as the cost of epigenetic profiling continues to fall, enabling longitudinal collection in larger population-representative studies and field experiments.

## Materials and Methods

### Data

#### The Health and Retirement Study (HRS)

The HRS is a nationally representative, biannual, longitudinal panel study of individuals over the age of 50 and their spouses that began in 1992. The study is sponsored by the National Institute on Aging (NIA U01AG009740) and is conducted by the University of Michigan (70). Comprehensive information about participants’ socioeconomic background, income, assets, and employment is collected from the time of respondent entry until death. The HRS introduces a new cohort of participants every six years and interviews around 20,000 participants every two years.

DNA methylation data were collected as part of the 2016 HRS Venous Blood Study (VBS). The DNAm sample is racially and socioeconomically diverse and representative of the full HRS sample (71). In sensitivity analyses, we adjusted for cell-type proportions using results from a white blood cell (WBC) differential assay (72). Demographic and socioeconomic data were taken from the RAND HRS Longitudinal File 2018 (V1) (73). Information on cause of death was taken from HRS exit interview files (74).

#### State-level measures of economic conditions

The following available measures were linked to HRS participants at the year of birth and state of birth levels: 1) *Wage Index (1929-1956):* farm and non-farm wages and salaries from the Bureau of Economic Analysis (BEA) (55); 2) *Employment Index (1929-1940):* employment in manufacturing and non-manufacturing sectors (57); 3) *Car Sales Index (1929-1940):* total number of car sales from the annual statistical issues of the industry trade publication, *Automotive Industries* (58). Measures were converted into indices by dividing the variable by its 1929 level and multiplied by 100 so each state has a value of 100 in 1929. We transformed the wage index to real wages using the consumer price index.

#### In utero exposure measure

Calendar year does not always correspond well with the prenatal period, however monthly state-level macroeconomic data were not available in the 1930s. To generate a more precise measure, we constructed a weighted average of in utero exposure as follows:

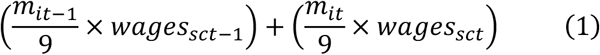

Where *m*_*it*−1_ and *m*_*it*_ reflect the approximate number of months individual *i* spent in utero in *t* − 1 and *t* relative to their month of birth, and values of the wage index are assigned according to *i*’s state (*s*) and year of birth (*c*). For example, an individual born in March received an in utero wage index equal to 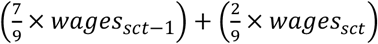. Since we do not know the exact number of months spent in utero, our exposure variable is still subject to measurement error; however, our results are larger in magnitude and more precise than in utero measures that correspond to quarter of birth (**SI Appendix, Table S16**).

#### Epigenetic aging measures

We used six different epigenetic clock or pace of aging measures that were constructed by the HRS from individual CpG-level data and are publicly available (75). Epigenetic age acceleration (EAA) was computed by using the residuals from regressions of each clock on chronological age. Residualization was not applied to DunedinPoAm since it already quantifies deviations in chronological age from the expected sample norm. **SI Appendix, Table S1** provides more details on the methods and number of CpG sites that were used to compute epigenetic aging measures according to author-specific algorithms.

#### Empirical Framework

Our baseline specification is as follows:

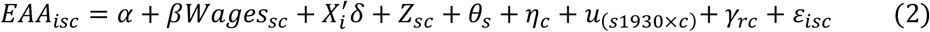

where *EAA*_*isc*_ is the epigenetic age acceleration outcome in 2016 for individual *i*, born in state *s* in year *c. Wages* represents the aggregate wage index at the state and year levels for the in utero period as defined in Equation (1). The matrix *X*_*i*_ contains individual characteristics at baseline including sex, race, and dichotomous indicators for maternal education (no degree, high school degree or more, and missing maternal education). *Z*_*sc*_ is a vector of state-level characteristics around 1930 interacted with year of birth, including the maternal (76) and infant mortality rate in 1928 (77), the population in 1930 (76), and whether the percent of farmland in a state was above the 75^th^ percentile nationally in 1930 (78). The term *u*_(*s*1930×*c*)_ represents a state’s share of wage earners in manufacturing in 1929 (79) interacted with year of birth fixed effects. We include these controls because the severity of cyclical fluctuations across states was driven in part by state differences in the proportion of manufacturing and agricultural industries (80). The terms *θ*_*s*_ and *η*_*c*_ are state and year of birth fixed effects, respectively. The geographic fixed effects help absorb time-invariant differences at the state level, while the time fixed effects absorb factors that vary over time but are invariant across states. To control for changes in regional conditions (e.g., economic or demographic) throughout the 1930s we include region-specific linear time trends, or the term *γ*_*rc*_. Robust standard errors are clustered at the state of birth level. All models were estimated using HRS sample weights for the 2016 VBS sample to adjust for sample composition.

Coefficients reported in **Figure 2** are from our second specification:

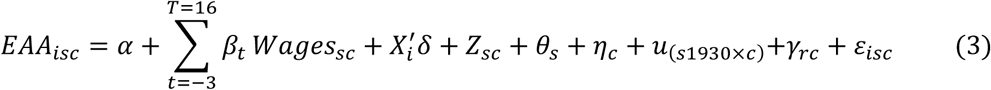

This model is identical to the baseline specification in Equation (2) except in addition to the in utero term, the model also conditions on a pre-trend for average wages in years *t* − 3 and *t* − 2 and eight additional two-year averaged terms for wage exposures when individuals were between the ages of one and sixteen.

## Data Availability

This study used restricted individual level information from the HRS and our contractual agreement does not permit public dissemination of the data. Details on how to access restricted data for the HRS can be found at https://hrs.isr.umich.edu/data-products/restricted-data. Publicly available data used in this study is referenced in the main text and the SI Appendix.

## Acknowledgements

The HRS is sponsored by the National Institute on Aging (NIA U01AG009740) and conducted by the University of Michigan. We thank Jason Fletcher, Dalton Conley, Dan Belsky, Josh Hausman, and David Cutler.

