## Supplementary Information for "In Utero Exposure to the Great Depression is Reflected in Late-Life Epigenetic Aging Signatures"

**This PDF file includes:**

Supplementary text
Figures S1 to S3
Tables S1 to S16
SI References

### 1. Overview of DNA methylation data and epigenetic aging measures

**Health and Retirement Study (HRS) DNA methylation data.** Detailed information on the 2016 Venous Blood Study (VBS) is provided in the VBS 2016 Data Description (1). Briefly, blood was collected from consenting respondents during in home phlebotomy visits. Every attempt was made to schedule the blood draw within four weeks of the 2016 HRS core interview. Fasting was recommended but not required. Methylation was measured using the Infinium Methylation EPIC BeadChip. Samples were randomized across plates by key demographic variables (i.e., age, cohort, sex, education, race/ethnicity) with 40 pairs of blinded duplicates. Analysis of duplicate samples showed a correlation >0.97 for all CpG sites. The *minfi* package in R software was used for data preprocessing, and quality control. 3.4% of the methylation probes (n=29,431 out of 866,091) were removed from the final dataset due to suboptimal performance (using a detection p-value threshold of 0.01). Analysis for detection p-value failed samples was done after removal of detection p-value failed probes. Using a 5% cut-off (*minfi*) 58 samples were removed. Sex mismatched samples and any controls (cell lines, blinded duplicates) were also removed. High quality methylation data was available for 97.9% samples (n=4,018). Missing beta methylation values were imputed with the mean beta methylation value of the given probe across all samples prior to constructing DNAm age measures.

**Epigenetic aging measures.** We used publicly available epigenetic aging measures that were constructed by the HRS from CpG level data (2). **Table S1** provides a basic overview of the six measures utilized in this study. We focused on first- and second-generation clocks and pace of aging measures that have been widely used in the literature and that were constructed using different methods, training phenotypes, and tissue samples.

**Table S1.** Characteristics of epigenetic aging clocks analyzed in the HRS

| Name | First author (year) | Training phenotype | Tissue type(s) used to derive clock | CpG sites (#) | Unit of measurement |
|---|---|---|---|---|---|
| Horvath | Horvath (2013) | Chronological age | 51 tissues/cells | 353 | Years |
| Hannum | Hannum (2013) | Chronological age | Whole blood | 71 | Years |
| SkinBlood | Horvath (2018) | Chronological age | Skin | 391 | Years |
| DNAmPhenoAge | Levine (2018) | Phenotypic age | Whole blood | 513 | Years |
| GrimAge | Lu (2019) | Mortality risk (time-to-death) | 7 plasma proteins, smoking pack years | 1,030 | Years |
| DunedinPoAm | Belsky (2020) | Rate of change across 18 biomarkers | Whole blood | 46 | Rate of aging in years |

First generation Horvath (3) and Hannum (4) clocks were developed using penalized regression methods (i.e., elastic net) to train a predictor of chronological age based on DNA methylation (DNAm) levels across the human genome that were captured by the Illumina Infinium arrays (27K or 450K). The **Horvath clock** was developed using approximately 8,000 samples from 82 datasets that incorporated 51 healthy tissues/cells to develop a multi-tissue predictor, or "pan-tissue" clock that includes 353 CpGs in its prediction. The **Hannum clock** was developed using whole blood samples from 656 persons aged 19-101 years and includes 71 CpG sites. The **SkinBlood clock** was developed using the 450K and EPIC platforms (850K) from human fibroblasts, keratinocytes, buccal cells, endothelial cells, lymphoblastoid cells, skin, blood, and saliva samples to provide greater prediction accuracy in fibroblasts and other cell types commonly used in ex vivo studies (e.g., blood or buccal swabs) (5). The 391 CpG sites in the clock had to have 1) high absolute correlation with chronological age in different cell types, or 2) little to no significant correlation with age. It predicts age for sorted neurons, glia, brain, liver, and bone samples, and was correlated with gestational age in cord blood.

Second generation clocks were trained on biomarkers of aging that reflect morbidity and/or mortality risk. The **DNAmPhenoAge clock** was developed using a two-step process from analysis in NHANES III and the InCHIANTI Study (6). In step one, a "phenotypic age" measure in units of years was developed using a Cox proportional elastic net model that trained nine biomarkers (albumin, creatinine, glucose, (log) C-reactive protein (CRP), lymphocyte percent, mean cell volume, red blood cell distribution width, alkaline phosphatase, and white blood cell count)) on chronological age. In step 2, the phenotypic age variable was used as the outcome for training an epigenetic clock in whole blood using elastic net (n=456) and CpG sites available on all three chips (27K, 450K, and EPIC). The DNAmPhenoAge clock has 513 CpG sites. The **GrimAge clock** was trained in the Framingham Heart Study Offspring Cohort using CpGs that are present on both the 450K and EPIC arrays (n=1,731) and was developed in two steps (7). In step one, DNAm-based surrogates of smoking pack-years and plasma proteins that have been previously associated with mortality or morbidity were defined using elastic net (adrenomedullin, CRP, plasminogen activation inhibitor 1 (PAI-1), and growth differentiation factor 15 (GDF15)). In step two, time-to-death was regressed on these DNAm-based surrogate biomarkers, age, and sex. The resulting mortality risk estimate from the regression model was then linearly transformed to be in units of years.

The pace of aging measure (**DunedinPoAm**) was developed in the Dunedin Study using the change in age-associated biomarkers over time among individuals who are the same chronological age (n=810) (8). In step one, the rate of change in 18 blood-chemistry and organ-system function biomarkers was modeled and then everyone's personal rate-of-change relative to the sample norm was calculated. All 18 personal rates of change were then combined into a composite for each Study member called the "Pace of Aging" (9). In step two, elastic net regression was then applied using Pace of Aging between ages 26 to 38 as the training outcome and all DNAm probes that appeared on both the 450K and EPIC arrays as potential predictor variables. DunedinPoAm has 46 CpG sites and it represents physiological decline experienced per one year of calendar time.

### 2. Summary statistics

**Table S2** displays summary statistics for the HRS analytic sample (n=832). Epigenetic age acceleration (EAA) measures were calculated by using the residuals from the regression of each clock on chronological age in the full Venous Blood Study (VBS) DNAm sample (n=4,018). Residualization was not applied to DunedinPoAm since it already quantifies deviations in chronological age from the expected sample norm. **Figure S1** displays correlations between DunedinPoAm and the EAA measures used in this study.

**Table S2.** Summary statistics for the HRS sample (n=832)

| | Mean | SD |
|---|---|---|
| **Exposures** | | |
| Wage index | 101.39 | 17.90 |
| Employment index | 92.24 | 12.58 |
| Car sales index | 62.27 | 21.98 |
| **Outcomes** | | |
| Horvath EAA | -0.13 | 7.07 |
| SkinBlood EAA | -0.003 | 4.82 |
| Hannum EAA | -0.05 | 5.80 |
| PhenoAge EAA | -0.08 | 6.94 |
| GrimAge EAA | 0.10 | 4.53 |
| DunedinPoAm | 1.08 | 0.09 |
| **Individual characteristics** | | |
| Birth year | 1937 | 2.54 |
| Chronological age | 79.03 | 2.61 |
| Female | 0.57 | 0.50 |
| White | 0.91 | 0.29 |
| Black | 0.08 | 0.27 |
| Other race | 0.01 | 0.11 |
| Hispanic | 0.03 | 0.16 |
| No degree | 0.15 | 0.36 |
| GED/HS degree | 0.58 | 0.49 |
| Associate's degree | 0.04 | 0.20 |
| Bachelor's degree | 0.22 | 0.42 |
| Mother's education=No degree | 0.51 | 0.50 |
| Mother's education=HS or higher | 0.43 | 0.50 |
| Mother's education missing | 0.05 | 0.23 |

*Note:* EAA=epigenetic age acceleration; SD=standard deviation. Statistics are weighted using HRS weights for the Venous Blood Study (VBS) sample.

**Figure S1**. Correlations between epigenetic age acceleration (EAA) and pace of aging measures in the HRS analysis sample (n=832)

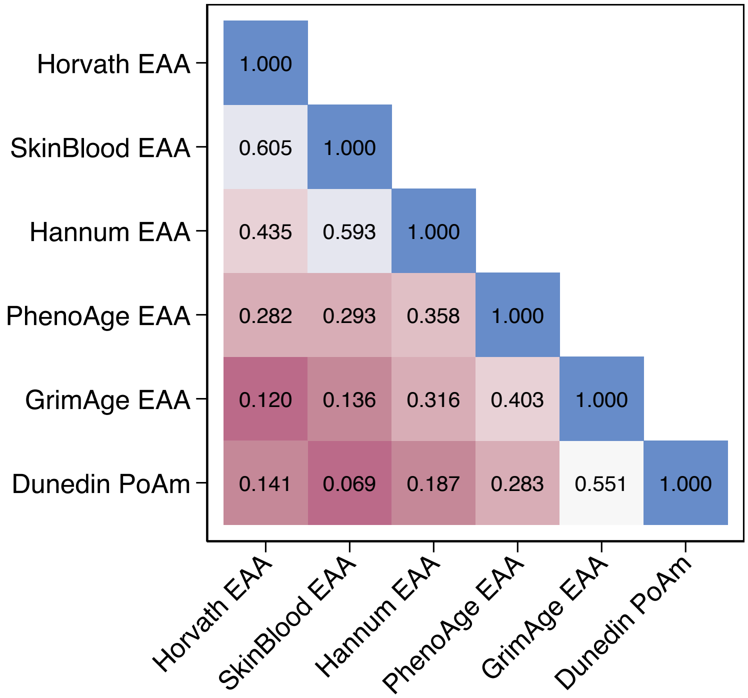

**Table S3.** Effect of the wage index in utero on GrimAge EAA and DunedinPoAm across empirical specifications

The table reports estimated effects of the wage index in utero on GrimAge EAA and DunedinPoAm from separate regressions that incrementally add baseline covariates and state- and regional-level controls. Regressions in Column (1) control for year of birth (YOB) and state of birth (SOB) fixed effects (FE). Column (2) adds individual covariates for sex and race. Column (3) adds controls for maternal education (no degree, high school and above, or missing). Column (4) adds the following additional state-level controls interacted with YOB linear time trends (LTT): infant mortality rate in 1928, the maternal mortality rate in 1929, and whether a state's share of farmland was in the 75$^{th}$ percentile nationally in 1930; Column (4) also includes indicators for whether state employment in manufacturing was in the 75$^{th}$ percentile nationally in 1929 times YOB FE. Column (5) adds region of birth controls interacted with YOB LTT (regions include New England, Middle Atlantic, East North Central, West North Central, South Atlantic, East South Central, West South Central, Mountain, and Pacific). All models were estimated using linear regression with VBS sample weights provided by the HRS. Robust standard errors clustered at the SOB level are in brackets.

| | **GrimAge EAA** | | | | | **DunedinPoAm** | | | | |
|---|---|---|---|---|---|---|---|---|---|---|
| | (1) | (2) | (3) | (4) | (5) | (1) | (2) | (3) | (4) | (5) |
| Wage index in utero | -0.0846*** | -0.0601** | -0.0662*** | -0.0873*** | -0.0950*** | -0.0013** | -0.0011** | -0.0012** | -0.0017** | -0.0022*** |
| | [0.0287] | [0.0231] | [0.0239] | [0.0301] | [0.0294] | [0.0005] | [0.0005] | [0.0005] | [0.0007] | [0.0007] |
| Observations | 832 | 832 | 832 | 832 | 832 | 832 | 832 | 832 | 832 | 832 |
| R-squared | 0.095 | 0.243 | 0.248 | 0.257 | 0.273 | 0.073 | 0.081 | 0.085 | 0.102 | 0.134 |
| YOB FE | X | X | X | X | X | X | X | X | X | X |
| SOB FE | X | X | X | X | X | X | X | X | X | X |
| Individual covariates | | X | X | X | X | | X | X | X | X |
| Maternal education | | | X | X | X | | | X | X | X |
| State-level controls*YOB | | | | X | X | | | | X | X |
| Share of manufacturing*YOB FE | | | | X | X | | | | X | X |
| Region of birth-specific LTT | | | | | X | | | | | X |

*Note:* YOB= year of birth; FE=fixed effect; LTT=linear time trend. *** p<0.01, ** p<0.05

### 3. Analyses with alternative state-level measures of economic conditions

**Employment index. Figure S2** depicts variation in the employment index across states, which includes employment in manufacturing and non-manufacturing sectors. Data were obtained from Wallis (1989) in index form (base year=1929) (10). The figure shows that the impact of the Depression was not spread equally across states. States in the South Atlantic region suffered smaller employment declines and experienced a more rapid recovery after 1933. This is attributed to differences in the composition of the labor force across states (the south had a higher concentration of industries that experienced smaller declines in employment), differential effects of institutional changes put in place by the New Deal, and the impact of the 1937-1938 monetary recession, which had a greater impact on states in the northeast (10).

**Figure S2.** Variation in the employment index across states, 1929-1940

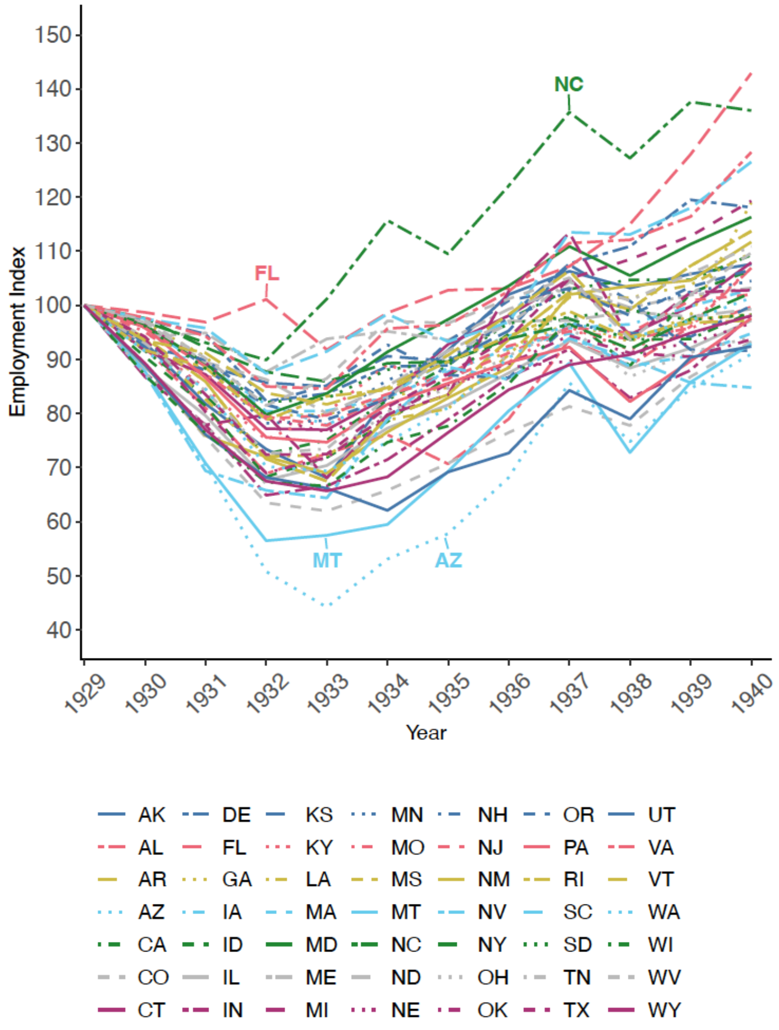

**Table S4.** Effect of the employment index on GrimAge EAA and DunedinPoAm

The table displays results from separate regressions of the effect of the employment index in the preconception, in utero, and postnatal years on GrimAge EAA or DunedinPoAm. Regressions in Column (1) were estimated in the same sample of individuals used in our preferred specification with the wage index and are therefore more directly comparable to the wage index results (n=832). To analyze effects in the postnatal period, results in Columns 2-3 were estimated in a subsample of these individuals (n=588) because state-level employment data from Wallis were not available after 1940. Results are from the fully specified model and include controls for SOB FE, YOB FE, sex, race, maternal education, YOB LTT for the infant mortality rate in 1928, the maternal mortality rate in 1929, and whether a state's share of farmland was in the 75$^{th}$ percentile nationally in 1930. Regressions also control for whether a state's employment in manufacturing was in the 75$^{th}$ percentile nationally in 1929 times YOB FE, and YOB LTT for region of birth. All models were estimated using linear regression with VBS sample weights provided by the HRS. Robust standard errors clustered at the SOB level are in brackets.

| | **GrimAge EAA** | | | **DunedinPoAm** | | |
|---|---|---|---|---|---|---|
| | (1) | (2) | (3) | (1) | (2) | (3) |
| Employment index ages -3 to -2 | -0.0195 | 0.0080 | 0.0064 | -0.0002 | -0.0007 | -0.0006 |
| | [0.0290] | [0.0452] | [0.0459] | [0.0010] | [0.0016] | [0.0016] |
| Employment index in utero | -0.1122*** | -0.0521 | -0.0518 | -0.0023*** | -0.0021** | -0.0022** |
| | [0.0360] | [0.0454] | [0.0452] | [0.0007] | [0.0010] | [0.0010] |
| Employment index ages 1 to 2 | | | -0.0086 | | | 0.0003 |
| | | | [0.0457] | | | [0.0010] |
| Observations | 832 | 588 | 588 | 832 | 588 | 588 |
| R-squared | 0.252 | 0.277 | 0.277 | 0.117 | 0.131 | 0.131 |

*Note:* *** p<0.01, ** p<0.05.

**Car sales index.** Data on car sales were collected by Hausman from annual statistical issues of the industry trade publication, *Automotive Industries* (11). Data on cars sales were used as a proxy for household consumption and have little measurement error because state laws mandated the registration of new cars (11). Thus, car sales have additional advantages as a macro indicator when compared to data on wages and employment. Data were converted into an index by dividing the variable by its 1929 level and multiplying by 100 so each state had a value of 100 in 1929. **Figure S3** depicts variation in the car sale index across states from 1929-1940. The figure shows that the U.S. experienced a second recession in 1938, adding to the large variation in economic conditions throughout the decade.

**Figure S3.** Variation in the car sale index across states, 1929-1940

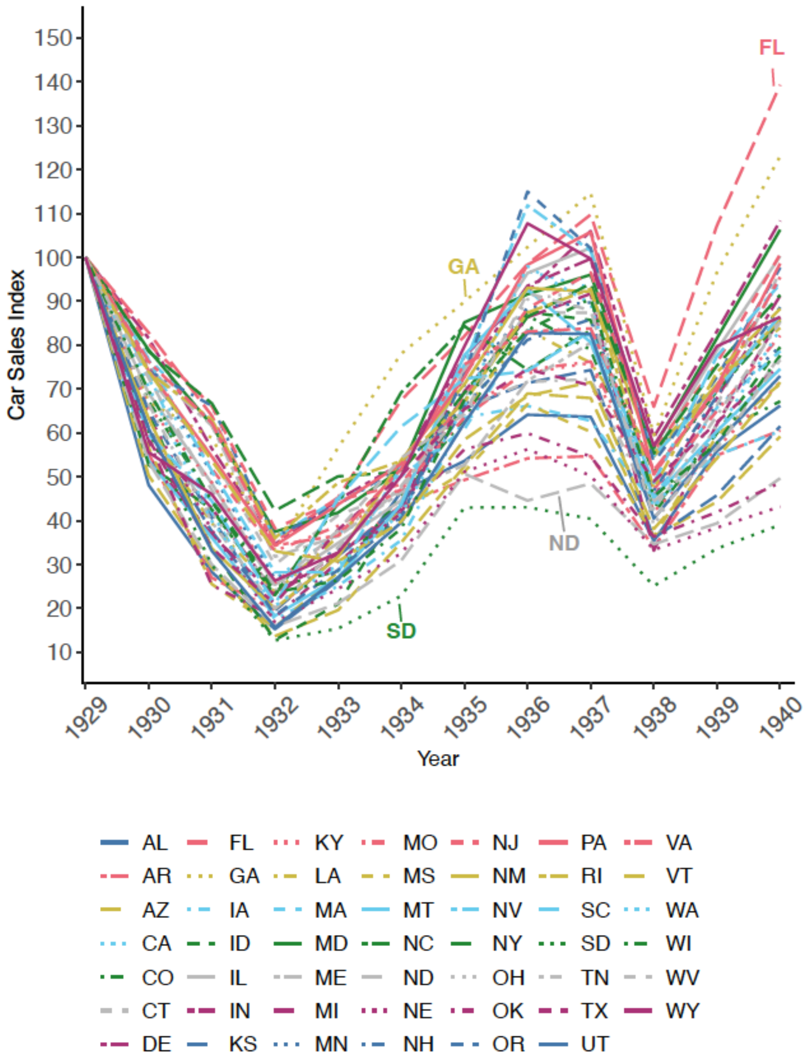

**Table S5.** Effect of the car sales index on GrimAge EAA and DunedinPoAm

The table displays results from separate regressions of the effect of the car sales index in the preconception, in utero, and postnatal years on GrimAge EAA or DunedinPoAm. Regressions in Column (1) were estimated in the same sample of individuals used in our preferred specification with the wage index and are therefore more directly comparable to the wage index results (n=832). To analyze effects in the postnatal period, results in Columns 2-3 were estimated in a subsample of these individuals because state-level car sale data from Hausman were not available after 1940. Results are from the fully specified model and include controls for SOB FE, YOB FE, sex, race, maternal education, YOB LTT for the infant mortality rate in 1928, the maternal mortality rate in 1929, and whether a state's share of farmland was in the 75th percentile nationally in 1930. Regressions also control for whether a state's employment in manufacturing was in the 75th percentile nationally in 1929 times YOB FE, and YOB LTT for region of birth. All models were estimated using linear regression with VBS sample weights provided by the HRS. Robust standard errors clustered at the SOB level are in brackets.

| | **GrimAge EAA** | | | **DunedinPoAm** | | |
|---|---|---|---|---|---|---|
| | (1) | (2) | (3) | (1) | (2) | (3) |
| Car sales index ages -3 to -2 | -0.0044 | 0.0180 | 0.0265 | -0.0003 | -0.0002 | -0.0003 |
| | [0.0247] | [0.0426] | [0.0417] | [0.0007] | [0.0012] | [0.0012] |
| Car sales index in utero | -0.0469*** | -0.0489*** | -0.0449** | -0.0011*** | -0.0011** | -0.0011** |
| | [0.0151] | [0.0174] | [0.0183] | [0.0004] | [0.0005] | [0.0005] |
| Car sales index ages 1 to 2 | | | 0.0528 | | | -0.0009 |
| | | | [0.0345] | | | [0.0010] |
| Observations | 832 | 588 | 588 | 832 | 588 | 588 |
| R-squared | 0.251 | 0.284 | 0.286 | 0.118 | 0.134 | 0.135 |

*Note:* *** p<0.01, ** p<0.05.

### 4. Additional sensitivity analyses

**Table S6.** Wage index estimates that account for white blood cell (WBC) proportions

The table reports the estimated effect of the wage index in utero on GrimAge EAA and DunedinPoAm after controlling for WBC proportions. Data on WBC proportions was assayed as part of the 2016 HRS Venous Blood Study (VBS) (n=9,934). Details on collection, participation, and quality control procedures are provided in the VBS 2016 Data Description (1). Regressions control for the proportion monocytes, lymphocytes, eosinophils, and basophils (omitted category=proportion neutrophils) and their interactions with YOB LTT. Coefficients are from the fully specified model and include controls for SOB FE, YOB FE, sex, race, maternal education, YOB LTT for the infant mortality rate in 1928, the maternal mortality rate in 1929, and whether a state's share of farmland was in the 75$^{th}$ percentile nationally in 1930. Regressions also control for whether a state's employment in manufacturing was in the 75$^{th}$ percentile nationally in 1929 times YOB FE, and YOB LTT for region of birth. All models were estimated using linear regression with VBS sample weights provided by the HRS. Robust standard errors clustered at the SOB level are in brackets.

| | GrimAge EAA | DunedinPoAm |
|---|---|---|
| Wage index in utero | -0.0691** | -0.0014* |
| | [0.0323] | [0.0007] |
| Proportion monocytes | 0.0846 | 0.0053* |
| | [0.1084] | [0.0030] |
| Proportion lymphocytes | -0.0326 | -0.0036*** |
| | [0.0450] | [0.0012] |
| Proportion eosinophils | -0.1495 | 0.0009 |
| | [0.1024] | [0.0032] |
| Proportion basophils | 0.2913 | 0.0042 |
| | [0.7784] | [0.0224] |
| Observations | 826 | 826 |
| R-squared | 0.370 | 0.332 |

*Note:* Omitted category=proportion neutrophils. Models also include cell proportions interacted with YOB LTT. *** p<0.01, ** p<0.05, * p<0.1.

**Table S7.** Wage index estimates that omit epigenetic aging outliers

Results in Table S7 omit individuals in our sample who are in the top and bottom 1% of the GrimAge EAA and DunedinPoAm distributions. Coefficients are from the fully specified model and include controls for SOB FE, YOB FE, sex, race, maternal education, YOB LTT for the infant mortality rate in 1928, the maternal mortality rate in 1929, and whether a state's share of farmland was in the 75$^{th}$ percentile nationally in 1930. Regressions also control for whether a state's employment in manufacturing was in the 75$^{th}$ percentile nationally in 1929 times YOB FE, and YOB LTT for region of birth. All models were estimated using linear regression with VBS sample weights provided by the HRS. Robust standard errors clustered at the SOB level are in brackets.

| | GrimAge EAA | DunedinPoAm |
|---|---|---|
| Wage index in utero | -0.0743** | -0.0023*** |
| | [0.0309] | [0.0006] |
| Observations | 816 | 816 |
| R-squared | 0.250 | 0.131 |

*Note:* *** p<0.01, ** p<0.05.

**Table S8.** Wage index estimates that account for population stratification

The table displays results from separate regressions estimated in individuals of European ancestry born between 1932 and 1940. Coefficients are from the fully specified model and include controls for the top ten principal components (PCs) of the European ancestry genetic data and their interactions with the wage index. PCs were obtained from the Social Science and Genetics Consortium Polygenic Index (PGI) Repository (12). Additional controls: SOB FE, YOB FE, sex, race, maternal education, YOB LTT for the infant mortality rate in 1928, the maternal mortality rate in 1929, and whether a state's share of farmland was in the 75th percentile nationally in 1930. Regressions also control for whether a state's employment in manufacturing was in the 75th percentile nationally in 1929 times YOB FE, and YOB LTT for region of birth. Models were estimated using linear regression with VBS sample weights provided by the HRS. Robust standard errors clustered at the SOB level are in brackets.

| | GrimAge EAA | DunedinPoAm |
|---|---|---|
| Wage index in utero | -0.1013*** | -0.0024*** |
| | [0.0316] | [0.0007] |
| Observations | 632 | 632 |
| R-squared | 0.302 | 0.190 |

*Note:* *** p<0.01

### 5. Impacts of other co-occurring historical events

**The Dust Bowl.** The Dust Bowl was an environmental catastrophe characterized by droughts, soil erosion, and severe dust storms that eroded sections of the Southern Plains and rendered millions of acres of formerly cultivated land useless for farming. Massive dust storms began in 1931 and persisted throughout the decade. Past studies have shown that cohorts exposed to this shock in childhood experienced significant declines in educational attainment and economic well-being in adulthood (13). Results that account for the Dust Bowl are consistent but suggest that the protective effect of higher state-level wages in utero was somewhat diminished for individuals who were simultaneously exposed to the Dust Bowl.

**Table S9.** Effect of the wage index in utero and being born in a Dust Bowl state on GrimAge EAA and DunedinPoAm

The table reports the estimated effect of the wage index on GrimAge EAA and DunedinPoAm with and without adjustments for being born in a Dust Bowl (DB) state. Results in Column (1) are from Table 1. Results in Column (2) include a dichotomous variable equal to one if an individual was born in a DB state and its interaction with the wage index. DB states included New Mexico, Colorado, Oklahoma, Kansas, and Texas. Additional controls: SOB FE, YOB FE, sex, race, maternal education, YOB LTT for the infant mortality rate in 1928, the maternal mortality rate in 1929, and whether a state's share of farmland was in the 75$^{th}$ percentile nationally in 1930. Regressions also control for whether a state's employment in manufacturing was in the 75$^{th}$ percentile nationally in 1929 times YOB FE, and YOB LTT for region of birth. Models were estimated using linear regression with VBS sample weights provided by the HRS. Robust standard errors clustered at the SOB level are in brackets.

| | **GrimAge EAA** | | **DunedinPoAm** | |
|---|---|---|---|---|
| | (1) | (2) | (1) | (2) |
| Wage index in utero | -0.0950*** [0.0294] | -0.0896*** [0.0291] | -0.0022*** [0.0007] | -0.0021*** [0.0007] |
| Wage index in utero*Born in a DB state | | -0.0530** [0.0209] | | -0.0007 [0.0007] |
| Observations | 832 | 832 | 832 | 832 |
| R-squared | 0.25 | 0.251 | 0.117 | 0.118 |

*Note:* DB=Dust Bowl. Models in Column 2 also control for the main effect of being born in a DB state. ***p<0.01, **p<0.05.

**The New Deal**. The Great Depression dramatically altered social spending in the U.S. In 1933, Roosevelt's New Deal programs began pouring funds into emergency work and disaster relief programs. Spending was more concentrated in areas with higher unemployment that were disproportionally affected by the economic downturn, and had far reaching impacts on socioeconomic and demographic outcomes, including income, employment, migration, mortality, crime rates, housing values, and home ownership rates (14–19). Results are consistent after adjusting for total state-level New Deal spending and its interaction with the wage index, which suggests that state-level variation in New Deal spending is not the primary driver behind our results with the wage index (**Table S10**).

**Table S10.** Effect of the wage index in utero and New Deal spending on GrimAge EAA and DunedinPoAm

The table reports the estimated effect of the wage index in utero on GrimAge EAA and DunedinPoAm with and without adjustments for New Deal spending and its interaction with the wage index. Results in Column (1) are from the preferred specification in Table 1. Results in Column (2) include a dichotomous variable for New Deal spending equal to one if an individual was born in a state that was in the top quartile of total per capita national spending in the 1930s and its interaction with the wage index. Additional controls: SOB FE, YOB FE, sex, race, maternal education, YOB LTT for the infant mortality rate in 1928, the maternal mortality rate in 1929, and whether a state's share of farmland was in the 75th percentile nationally in 1930. Regressions also control for whether a state's employment in manufacturing was in the 75th percentile nationally in 1929 times YOB FE, and YOB LTT for region of birth. Models were estimated using linear regression with VBS sample weights provided by the HRS. Robust standard errors clustered at the SOB level are in brackets.

| | **GrimAge EAA** | | **DunedinPoAm** | |
|---|---|---|---|---|
| | (1) | (2) | (1) | (2) |
| Wage index in utero | -0.0950***<br>[0.0294] | -0.0885***<br>[0.0301] | -0.0022***<br>[0.0007] | -0.0023***<br>[0.0007] |
| Wage index in utero*New Deal spending | | -0.0346*<br>[0.0196] | | 0.0003<br>[0.0005] |
| Observations | 832 | 832 | 832 | 832 |
| R-squared | 0.25 | 0.251 | 0.117 | 0.117 |

*Note:* Models in Column 2 also control for the main effect of New Deal spending. ***p<0.01, *p<0.1.

**Data on New Deal spending** are from Fishback, Kantor, and Wallis (20). To generate per capita state estimates, we aggregated data from the county level to the state level and divided by the state population in 1930. Following Fishback et al. (15), total spending included spending on relief (Federal Emergency Relief Administration grants, Civil Works Administration grants, Works Progress Administration grants, Public Assistance grants—Social Security Act), public works (Public Works Administration federal and nonfederal grants and nonfederal loans, public roads administration grants, public buildings administration grants), farm programs (Agricultural Adjustment Administration grants, Farm Credit Administration loans, Farm Security Administration Rural Rehab grants, Farm Security Administration Rural Rehab loans, Rural Electrification Administration loans), and the housing market (Reconstruction Finance Corporation loans, Home Owners Loan Corporation loans, U.S. Housing Administration loan contracts, U.S. Housing Administration grants).

**World War II (WWII).** Lastly, we examined whether our estimates are sensitive to WWII mobilization rates. The war not only represented a large-scale induction of men into the armed forces but also drew many women into the labor force, both of which may have had consequences on the stability of home and family life for children born in the 1930s (21, 22). To examine the impact of WWII mobilization on our results, we used data on average mobilization rates compiled by Acemoglu, Autor, and Lyle (23) and interacted mobilization rates with the wage index. Results are robust to adjustments for above average WWII mobilization rates across states and their interaction with the wage index, indicating our results are likely not influenced by downstream disruptions from WWII (**Table S11**).

**Table S11.** Effect of wage index in utero and WWII mobilization rates on GrimAge EAA and DunedinPoAm

The table reports the estimated effect of the wage index in utero on GrimAge EAA and DunedinPoAm with and without adjustments for WWII mobilization and its interaction with the wage index. Results in Column (1) are from the preferred specification in Table 1. Results in Column (2) include a dichotomous variable for WWII mobilization equal to one if an individual was born in a state that had a WWII mobilization rate that was above the national average, or a high mobilization rate (HMR), and its interaction with the wage index. Additional controls: SOB FE, YOB FE, sex, race, maternal education, YOB LTT for the infant mortality rate in 1928, the maternal mortality rate in 1929, and whether a state's share of farmland was in the 75th percentile nationally in 1930. Regressions also control for whether a state's employment in manufacturing was in the 75th percentile nationally in 1929 times YOB FE, and YOB LTT for region of birth. Models were estimated using linear regression with VBS sample weights provided by the HRS. Robust standard errors clustered at the SOB level are in brackets.

| | **GrimAge EAA** | | **DunedinPoAm** | |
|---|---|---|---|---|
| | (1) | (2) | (1) | (2) |
| Wage index in utero | -0.0950***<br>[0.0294] | -0.09373***<br>[0.0298] | -0.0022***<br>[0.0007] | -0.0023***<br>[0.0007] |
| Wage index in utero*HMR | | -0.0126<br>[0.0182] | | 0.0003<br>[0.0006] |
| Observations | 832 | 832 | 832 | 832 |
| R-squared | 0.25 | 0.25 | 0.117 | 0.117 |

*Note:* Models in Column 2 also control for the main effect of HMR. ***p<0.01.

### 6. Mortality analysis

**Table S12.** Effect of wages in utero and maternal education on the probability of survival

The table reports the estimated effect of the wage index in utero on the probability of survival until age 65, 75, and 85. Column (1) reports results from separate regressions that estimate the relationship between the wage index in utero and the probability of survival using all individuals in the HRS who were born between 1929 and 1940 (n=7,898). Results were calculated using all information on mortality through the 2018 HRS wave and were estimated using a linear probability model. Models in Column (2) add an interaction between the wage index and a dichotomous variable equal to one if a respondent reports that their mother did not have a high school degree (< 12 years of education) or if information on maternal education was missing. All models control for SOB FE, YOB FE, sex, and race and were estimated using sample weights for the HRS sample provided by the HRS. Robust standard errors clustered at the SOB level are in brackets.

| | (1) | (2) |
|---|---|---|
| **Probability of survival to age 65** | | |
| Wages in utero | -0.00006<br>[0.0004] | 0.00001<br>[0.0004] |
| Wages in utero*Mother did not have a degree | | -0.00004<br>[0.0003] |
| N | 7,898 | 7,898 |
| **Probability of survival to age 75** | | |
| Wages in utero | 0.0017**<br>[0.0007] | 0.0026***<br>[0.0008] |
| Wages in utero*Mother did not have a degree | | -0.0010*<br>[0.0006] |
| N | 7,898 | 7,898 |
| **Probability of survival to age 85** | | |
| Wages in utero | 0.0022**<br>[0.0008] | 0.0034***<br>[0.001] |
| Wages in utero*Mother did not have a degree | | -0.0014*<br>[0.0007] |
| N | 7,898 | 7,898 |

*Note:* Models also control for the main effect of low or missing maternal education
*** p<0.01, ** p<0.05.

**Table S13.** Effect of wages in utero on cause of death probabilities

The table reports results from separate regressions that estimate the relationship between the wage index in utero and the cause of death for all individuals in the HRS who were born between 1929 and 1940 (n=2,140). Results were calculated using all information on mortality through the 2018 HRS wave and were estimated using a linear probability model. “Heart condition” includes deaths related to heart, circulatory, and blood conditions; “metabolic conditions” includes deaths related to endocrine, metabolic, and nutritional conditions; “digestive system” includes deaths related to stomach, liver, gallbladder, kidney, and bladder conditions. All models control for SOB FE, YOB FE, sex, race, and maternal education and were estimated using sample weights for the HRS sample provided by the HRS. Robust standard errors clustered at the SOB level are in brackets.

| | Heart condition | Metabolic conditions | Digestive system | Neurological/ sensory condition | Emotional/ psychological condition |
|---|---|---|---|---|---|
| Wage index in utero | 0.0011<br>[0.0024] | -0.0018**<br>[0.0009] | 0.0006<br>[0.0009] | -0.0008<br>[0.0008] | 0.00001<br>[0.0001] |
| Observations | 2,140 | 2,140 | 2,140 | 2,140 | 2,140 |
| R-squared | 0.039 | 0.039 | 0.042 | 0.041 | 0.043 |

*Note:* ** p<0.05.

### 7. Inverse probability weighted (IPW) estimates

To construct inverse probability weights for mortality selection, we first estimated the probability of survival into our sample using a probit regression:

$$\Pr(Survival_{isc}) = \Phi\left(\alpha + \beta Wages_{sc} + X_i'\delta + \theta_s + \eta_c\right),$$

where the outcome was either survival until age 75 (the age that we first observe mortality selection in our HRS sample) or survival until 2016 (the year epigenetics were profiled in the HRS). *Wages* represents the aggregate wage index at the state and year levels for the in utero period as defined in Equation (1) in the main text. Results were estimated using all individuals in the HRS who were born between 1929 and 1940 (n=8,112 for survival until 2016, n=9,190 for survival until age 75). The matrix $X_i$ contains individual characteristics at baseline including sex and race; models were also run with controls for maternal education (dichotomous variable equal to one if the respondent's mother had less than 12 years of education or education status was missing and zero otherwise) and its interaction with the wage index. $\theta_s$ and $\eta_c$ are state and year of birth fixed effects, respectively. Robust standard errors were clustered at the state of birth level.

Results from probit regressions were used to estimate the inverse fitted probability of survival $\left(\frac{1}{\Pr(Survival_{isc})}\right)$ for survival until age 75 or survival until 2016. These were then used as probability weights to weight regression estimates in Table 2 so they are more reflective of the sample prior to mortality selection (24–26). In contrast to the published longitudinal weights that are supplied by the HRS, these weights were specifically designed for the problem of mortality-related non-response in our sample. Results from IPW estimation in Table 2 are similar to estimates in Table 1, which provides support against mortality-selection-related bias (26).

Finally, due to established differences in age-related mortality across demographic groups, we also created probability weights from probit regressions that were stratified by self-reported race and sex as a robustness check. Results from IPW estimation were unchanged when we applied weights estimated from race- and sex-specific survival analyses.

### 8. Other aging outcomes and future mortality

**Table S14.** Effect of the wage index in utero on other aging outcomes

The table reports the estimated effect of the wage index in utero on four aging outcomes. The frailty index (27) is equal to 1 if individuals report at least 1 of the following 5 conditions: wasting (loss of 10% or more of their body weight over a 2-year period); weakness (difficulty lifting/carrying weights over 10lbs because of health problems); slowness (difficulty getting up out of a chair after sitting for long periods because of health problems); fatigue or exhaustion; or a fall in the last 2 years. Metabolic syndrome index is the average of four mean standardized outcomes: doctor-diagnosed diabetes mellitus, coronary heart disease, stroke, and high blood pressure. Self-reported health status was rated on a five-point scale (1=excellent, 2=very good, 3=good, 4=fair, or 5=poor). Number of chronic conditions is the total count of the following doctor diagnosed disease conditions ever reported by the respondent: high blood pressure, diabetes mellitus, any type of cancer (except minor skin cancers), chronic lung disease, coronary heart disease, stroke, arthritis, or psychiatric problems. Additional controls: SOB FE, YOB FE, sex, race, maternal education, YOB LTT for the infant mortality rate in 1928, the maternal mortality rate in 1929, and whether a state's share of farmland was in the 75$^{th}$ percentile nationally in 1930. Regressions also control for whether a state's employment in manufacturing was in the 75$^{th}$ percentile nationally in 1929 times YOB FE, and YOB LTT for region of birth. Models were estimated using linear regression with HRS sample weights. Robust standard errors clustered at the state of birth level in brackets.

| | Frailty Index | Metabolic Syndrome Index | Self-Reported Health Status | Number of Chronic Disease Conditions |
|---|---|---|---|---|
| Wage index in utero | -0.0066 | -0.0158* | -0.0116 | -0.0312** |
| | [0.0044] | [0.0093] | [0.0089] | [0.0123] |
| Observations | 832 | 832 | 832 | 832 |
| R-squared | 0.146 | 0.133 | 0.143 | 0.135 |
| Mean | 0.772 | 0.394 | 2.903 | 3.341 |
| Effect size per 1 SD increase in the wage index | 0.281 SD | 0.275 SD | 0.203 SD | 0.37 SD |

*Note:* **p<0.05; *p<0.1.

**Table S15.** Association between aging measures in 2016 and the probability of death in 2018

The table reports estimated effects of six different aging measures in 2016 on the probability of death in 2018 (outcome). Results were estimated using linear probability models. Models include controls for YOB FE, SOB FE, race, sex, and dichotomous indicators for maternal education (no degree, high school degree and above, or missing). Robust standard errors clustered at the state of birth level in brackets.

| | Outcome: Probability of death in 2018 | | | | | |
|---|---|---|---|---|---|---|
| | (1) | (2) | (3) | (4) | (5) | (6) |
| GrimAge EAA | 0.0100***<br>[0.0023] | | | | | |
| DunedinPoAm | | 0.3008**<br>[0.1193] | | | | |
| Metabolic syndrome index | | | 0.0145<br>[0.011] | | | |
| Frailty index | | | | 0.0383*<br>[0.0204] | | |
| Self-reported health status | | | | | 0.0394***<br>[0.0116] | |
| Number of chronic disease conditions | | | | | | 0.0143**<br>[0.007] |
| Observations | 832 | 832 | 832 | 832 | 832 | 832 |
| R-squared | 0.113 | 0.099 | 0.091 | 0.093 | 0.111 | 0.095 |
| Outcome mean | 0.073 | | | | | |
| Effect size per 1 SD increase in aging measure | 0.173 SD | 0.101 SD | 0.057 SD | 0.061 SD | 0.154 SD | 0.077 SD |

*Note:* ***p<0.01, **p<0.05, *p<0.1.

**Table S16.** Effect of the wage index using alternative measures for the in utero period

The table reports estimated effects of the wage index in utero on GrimAge EAA and DunedinPoAm using two different measures of the in utero time period (M1 and M2). M1 is the wage index in utero variable used in this study (weighted average based on month of birth). The M2 in utero measure assigns the wage index in the year prior to birth if an individual was born in the first or second quarter of the year and assigns the wage index in the year of birth if an individual was born in the third or fourth quarter of the year (28). Additional controls: SOB FE, YOB FE, sex, race, maternal education, YOB LTT for the infant mortality rate in 1928, the maternal mortality rate in 1929, and whether a state's share of farmland was in the 75$^{th}$ percentile nationally in 1930. Regressions also control for whether a state's employment in manufacturing was in the 75$^{th}$ percentile nationally in 1929 times YOB FE, and YOB LTT for region of birth. Models were estimated using linear regression with VBS sample weights provided by the HRS. Robust standard errors clustered at the state of birth level in brackets.

| | GrimAge EAA | | DunedinPoAm | |
|---|---|---|---|---|
| | (1) | (2) | (1) | (2) |
| Wage index in utero (M1) | -0.0950***<br>[0.0294] | | -0.0022***<br>[0.0007] | |
| Wage index in utero (M2) | | -0.0540**<br>[0.0255] | | -0.0012**<br>[0.0006] |
| Observations | 832 | 832 | 832 | 832 |
| R-squared | 0.25 | 0.247 | 0.117 | 0.112 |

*Note:* ***p<0.01, **p<0.05.
